# Public opinion on global distribution of COVID-19 vaccines: evidence from two nationally representative surveys in Germany and the United States

**DOI:** 10.1101/2021.08.16.21262116

**Authors:** Matthias Klumpp, Ida G. Monfared, Sebastian Vollmer

**Author notes:** Corresponding author: Ida G. Monfared. **Author Contributions:** All authors contributed equally and order is following the alphabet. **Competing Interest Statement:** Authors declare that they have no competing interests.

## Abstract

**Background:** Despite ongoing calls for more equity in the global distribution of COVID-19 vaccines, there remains a great disparity between high- and low- or middle-income countries. Based on the principles of distributive justice, we assessed the public opinion on this issue in the United States and Germany as examples for high-income countries with a high potential for redistribution.

**Methods:** We conducted representative surveys among the adult population in the United States (*N*=1,000) and Germany (*N*=1,003) in June 2021 using two instances of an analytic hierarchy process (AHP) to elicit how the public weighs different principles and criteria according to which the vaccines should be allocated as well as discrete choice experiments to split a limited supply of vaccine doses between a hypothetical high-income and low-income country.

**Findings:** In the first AHP, respondents in the United States and Germany gave weight to “medical urgency” by 37·4% (37·2-37·5) and 49·4% (49·2-49·5), “equal access for all” 32·7% (32·6-32·8) and 25·4% (25·2-25·5), “production contribution” 13·7% (13·6-13·8) and 13·3% (13·2-13·4), and “free market rules” 16·3% (16·2-16·4) and 12·0% (11·9-12·1), respectively. In the discrete choice experiment responds in the United States split available vaccine doses such that the low-income country on average received 53·9 percent (95% CI: 52·6-55·1). For Germany this number was 57·5 percent (95% CI: 56·3-58·7). The low-income country had three times as many inhabitants as the high-income country. When facing the dilemma where a vulnerable family member was waiting for a vaccine as opposed to when there was no clear self-interest, 20·7% (18·2-23·3) of respondents in the United States and 18·2% (15·8-20·6) in Germany reduced the amount they allocated to the low-income country

**Interpretations:** The public in the United States and Germany favours utilitarian and egalitarian distribution principles of vaccines for COVID-19 over the currently prevailing libertarian or meritocratic principles. This implies that political approaches and decision favouring higher levels of redistribution would be supported by the public opinion in these two countries.

**Funding:** German Research Foundation DFG RTG 1723.

## Background

For the first time in history, a vaccine was developed during a global pandemic within a 12-months timeframe. Vaccine production resources are scarce and do not match demand – and this squeeze will continue until at least 2022 (1). The situation regarding virus variants and possible refreshing shots due to declining vaccination protection over time make global (re-)distribution decisions between countries even more complicated (2). These special circumstances bring about new questions regarding distributive justice and rationale of scarce COVID-19 vaccine doses on a global scale (3, 4). There are great disparities between countries across the globe with respect to access to COVID-19 vaccine doses. As of mid October 2021, only 2.8% of people in low-income countries have received at least one dose of a COVID-19 vaccine (5). Meanwhile and by the same date, 65.2% of people in the United States and 68.3% in Germany have received at least one dose of the vaccine and both countries have secured a vaccine supply that would suffice 337% and 242% of their population, respectively (6). The emergence of new variants shows that people in high-income countries (HICs) are not safe although fully vaccinated unless actions are taken globally (7). Despite several calls for equity in global distribution of vaccines (8, 9), the COVID-19 Vaccine Global Access program (COVAX) remains far behind its target (10). The reluctance of countries who could afford to donate more vaccines might be due to the unpredictable nature of the pandemic but in addition it could reflect political concerns about public disagreement. This raises the question which distribution principles are favoured by the population in the HICs that have ample access to vaccines: (i) distributing the vaccines according to greater outcome (utilitarian), (ii) equality for all (egalitarian as intended by COVAX), (iii) merits based on R&D or vaccine production efforts (11), or the present status quo, (iv) that is countries who pre-ordered or having higher purchasing power (free-market/libertarian) (12) keeping their advantage? In this line of thought, our study investigates which of these grounds are supported by the populations in the United States and Germany as the leading countries in R&D of COVID-19 vaccines. Moreover, these countries were particularly selected as they have the capacity to share COVID-19 vaccines with LMICs. Early studies conducted in the United States in July 2020 (13) and in Nov.-Dec. 2020 (14), respectively found that 39.6% and 22% of respondents were willing to donate more than 10% of the country’s vaccine supply to the WHO for distribution across those countries which have insufficient resources to buy their own. However, this was before a devastating winter wave with a large number of daily deaths in the United States and Germany being reported in Jan.-Feb. 2021 (15). Moreover, it is plausible that when it comes to own or a loved one’s life, people change their level of generosity in terms of sharing a lifesaving resource as opposed to when there is no clear case of self-interest (16, 17).

## Method

The study instrument (Appendix A, section II) consisted of four main sections. In the first module through Analytical Hierarchy Process (AHP), respondents were asked when it came to fair distribution of COVID-19 vaccines across the world which ground they believed to be more important. These grounds were “equal access for all”, “medical urgency”, “free market rules apply”, and “production contribution” representing egalitarian, utilitarian, libertarian, and merits. Respondents were presented with combinations of each pair in random order.

The second module was a discrete choice experiment asking respondents to divide a hypothetical 100 million doses of vaccines between two countries A and B. These two countries resemble a high- and low-income country, e.g. country B had 300 million inhabitants, 3,000 COVID-19 deaths per day, and ordered 100 million vaccine doses while country A had 100 million inhabitants, 200 COVID-19 deaths per day, and ordered 1,000 million doses of vaccines (Appendix A, section II, questions 6-8). This experiment was first carried out under the veil of ignorance. Then respondents were asked to think of a vulnerable family member being on the waiting list in country A with the information that this person’s place on the weighting was equal to the amount allocated to country A in the previous scenario plus 10 million. This meant that for this person to receive the vaccine, the respondent had to reduce the amount given to country B by at least 10 million. In the third scenario, respondents were asked to play the same allocation game but this time, consider themselves to be on the waiting list in a place where they needed to add 30 million additional doses to the amount given to country A to receive the vaccine themselves. These changes of scenarios added an element of self-interest at a time when many respondents were still waiting for their first dose of the vaccine.

In a third module, respondents were asked for their level of agreement (on a 7-point Likert-type scale, 1 = *Strongly disagree* to 7 = *Strongly agree*) to wait for their own vaccine for 3 months so that people in countries with specific characteristics could be vaccinated earlier. These characteristics which were individually presented were being more populated, higher number of COVID-19 deaths, lower number of intensive care hospital beds, a higher number of vaccine pre-orders, a higher annual income per head, investment in research and development of the vaccines, and production capacity for vaccines.

The fourth module was a second AHP asking respondents to rate the importance of seven criteria for vaccine distribution that where “number of inhabitants”, “number of daily COVID-19 deaths”, “number of intensive care unit (ICU) beds per 100 k population”, “number of vaccines pre-ordered”, “annual income per head (GDP)”, “investment in vaccine research and development (R&D)”, and “vaccine production capacity”.

To control for potential biases, respondents were asked whether or not they have already been vaccinated, to what extend COVID-19 had a negative impact on their lives (7-point Likert-type scale, 1 = *Strongly disagree* to 7 = *Strongly agree*), and also regarding their tendency for social desirability bias using the short 5-item version of the Marlowe-Crowne scale (SD5) (18). For the discrete choice experiment, we used a simple logistic regression model to investigate potential correlations between respondents’ characteristics and their decision to reduce the amount allocated to the low-income country when facing a scenario with a clear self-interest.

A minimum sample size of 1,000 was set following the method used by Gallup for conducting national polls (19). Using quotas, the sample was drawn to be representative of the adult population (aged 18 and above) in the United States and Germany according to age, gender, education, and state. Quotas were also placed for the vaccination rate to be representative of the general public at time of the fieldwork, from 25 May 2021 to 26 June 2021. Participants were recruited by a professional survey agency (INFO located in Berlin). Overall, 1,000 responses in the US and 1,003 in Germany were collected (for respondents’ demographics see Appendix A, Table A1). AHP data that were clearly unreliable (respondents selected the same response options, e.g., the first one, for all the possible comparisons) or inconsistent (AHP inconsistency index indicating randomness) were removed from the dataset. AHP weighting was calculated by using the dashboard Expert Choice Comparion and Stata 14.2 was used for the statistical analyses.

## Results

In the first AHP model where the four principles were compared, “medical urgency” received 37·4% (95% CI: 37·2-37·5) and 49·4% (95% CI: 49·2-49·5) of the United States and German participants weighting, respectively· This was followed by “equal access for all” receiving 32·7% (95% CI: 32·6-32·8) of the United States and 25·4% 25·4 (95% CI: 25·2-25·5) of the German respondents’ overall weighting across the four principles. “Production contribution” received 13·7% (95% CI: 13·6-13·8) and 13·3% (95% CI: 13·2-13·4) and “free market rules apply” 16·3% (95% CI: 16·2-16·4) and 12·0% (95% CI: 11·9-12·1) weighting in the United States and Germany, respectively.

Table 1 presents the characteristics of respondents who gave the highest weight to a particular principle. Respondents who gave the highest weight to a certain criterion do not systematically differ from respondents who gave the highest weight to another criterion by age, gender, education level or employment status.

**Table 1.** Characteristics of respondents who gave the highest weight to a particular criterion (in %; 95% CI in parentheses)

| US | Medical urgency | Equal access for all | Free market rules apply | Production contribution |
| --- | --- | --- | --- | --- |
| Age group >= 70 | 13 (9.5-16.4) | 16.7 (12.3-21) | 11.9 (4.6-19.2) | 16.2 (10.4-22) |
| Female | 51.5 (46.4-56.6) | 55.9 (50.1-61.7) | 40.1 (28.9-51.2) | 49.2 (41.3-57) |
| Education: bachelor or higher | 31.6 (26.8-36.4) | 30.8 (25.4-36.2) | 22.2 (12.8-31.7) | 30.9 (23.6-38.1) |
| Employment: employed (any type) | 50.9 (45.7-56) | 44.3 (38.5-50.1) | 56.1 (44.9-67.4) | 53.4 (45.5-61.2) |
| **DE** |  |  |  |  |
| Age group >= 70 | 16.9 (13.8-20) | 15.2 (9.5-21) | 19.8 (6.9-32.7) | 25.3 (17.9-32.8) |
| Female | 51 (46.9-55.1) | 54.3 (46.3-62.3) | 58.3 (42.3-74.3) | 47.9 (39.4-56.5) |
| Education: Abitur or technical diploma | 35.9 (32-39.9) | 26.5 (19.4-33.6) | 26.4 (12.2-40.7) | 23.3 (16.1-30.6) |
| Employment: employed (any type) | 58.7 (54.7-62.8) | 57.2 (49.3-65.1) | 48.3 (32.1-64.4) | 51.4 (42.9-60) |

In the second AHP, we replaced the abstract principles from the first AHP with seven concrete criteria as their proxies. Respondents in the United States and Germany gave the following weightings to each criteria with 95% CI presented in parentheses: “number of inhabitants” 14·4% (14·4-14·5) and 12·9% (12·8-12·9), “number of daily COVID-19 deaths” 28·7% (28·6-28·8) and 32% (31·9-32·1), “number of intensive care hospital beds (per 100,000)” 18·6% (18·5-18·6) and 23% (22·9-23), “number of vaccines pre-ordered” 10·4% (10·3-10·4) and 9·3% (9·3-9·4), “annual income per head (GDP)” 5·2% (5·2-5·2) and 4·4% (4·4-4·4), “invested in vaccine research and development” 10·8% (10·7-10·8) and 8·9% (8·8-8·9), and “vaccine production capacity” 11·9% (11·9-12) and 9·5% (9·5-9·6). Figure 1 presents how the first and second AHP results compare with each other.

**Figure 1.**
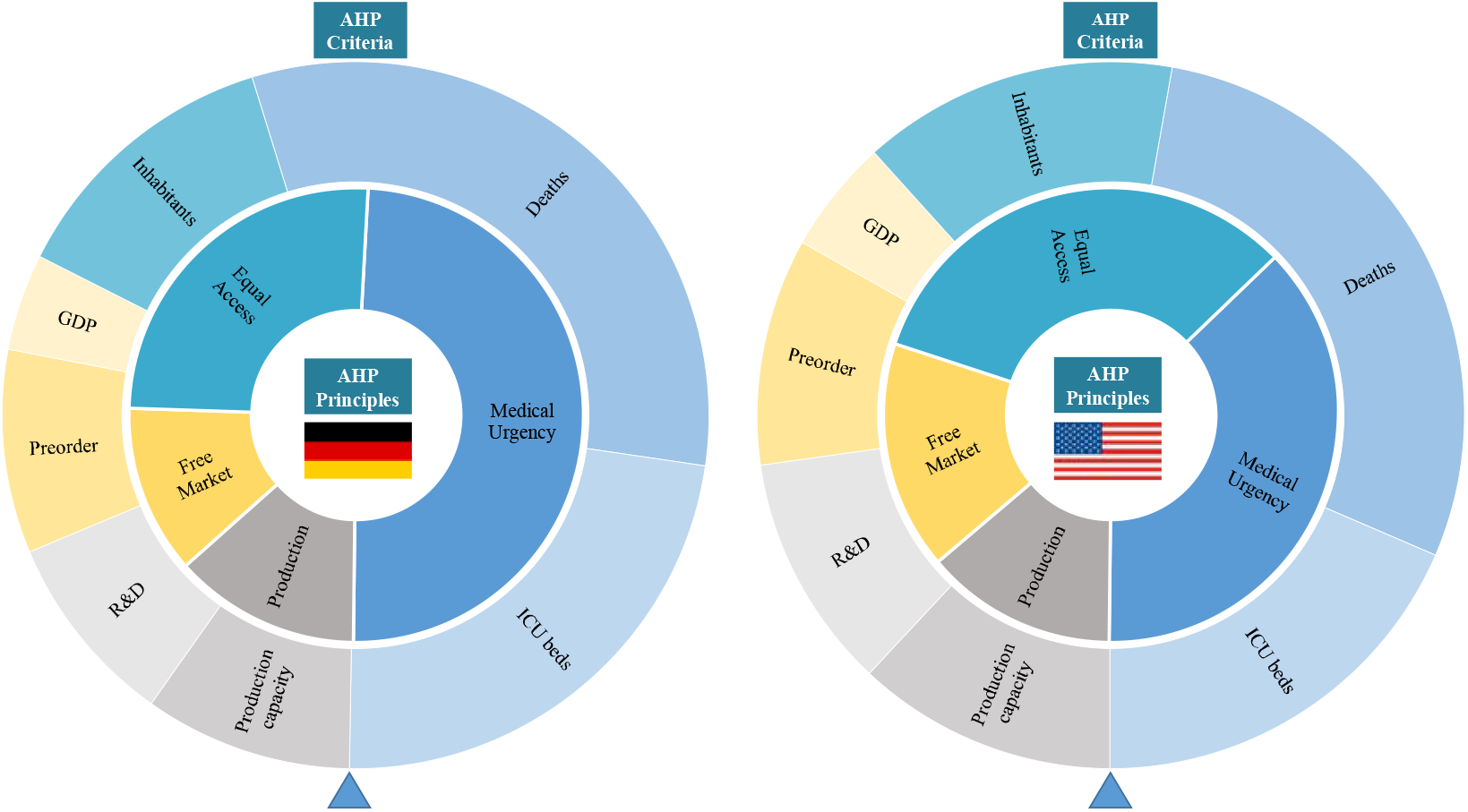
Weighting of 4 principles (inner circle) versus 7 criteria (outer circle).

In the experiment during which participants were asked to divide 100 million doses of vaccines between countries A and B, in a general scenario and when there was no clear self-interest, on average, respondents in the United States gave 53·9 million (95% CI: 52·6-55·1) and in Germany gave 57·5 million (95% CI: 56·3-58·7) of available doses to country B, which had three times as many inhabitants as country A. When they were faced with a situation where one of their vulnerable family members was on the waiting list, 20·7% (95% CI: 18·2-23·3) of respondents in the United States and 18·2% (95% CI: 15·8-20·6) in Germany reduced the amount they have given to country B by at least 10 million doses (the minimum amount needed to be added to the share of country A so that their family member could receive a vaccines). In the third scenario and when they were asked what if they themselves were on the waiting list, 6·9% (95% CI: 5·3-8·5) of respondents in the United States and 5·7% (95% CI: 4·3-7·2) in Germany reduced the amount allocated to country B by at least 30 million doses (the minimum amount they needed to add for country A allocation so that they themselves would receive a vaccine as opposed to when the scenario was rather general).

Table 2 presents the results of a logit regression with a binary variable as outcome that a responded reduced the amount allocated to country B by enough to have sufficient supply for a vulnerable family member or the respondents themselves. We do not find a significant association of this decision with gender, education level, employment status, vaccination status or previous exposure to COVID-19. There was an association with an age of 70 and above which was statistically significant for the scenario with a vulnerable family member in Germany.

**Table 2.** Odds ratio (*R*) of reducing the amount given to country B enough to have sufficient supply for a vulnerable family member or the respondents themselves.

|  | Vulnerable family member |  | Oneself |  |
| --- | --- | --- | --- | --- |
|  | US | DE | US | DE |
|  | (1) *OR* | (2) *OR* | (3) *OR* | (4) *OR* |
| Age group >= 70 | 1.49<br>(0.93 - 2.38) | 1.81**<br>(1.08 - 3.02) | 1.41<br>(0.69 - 2.89) | 1.66<br>(0.79 - 3.48) |
| Female = 1 | 0.99<br>(0.71 - 1.39) | 1.15<br>(0.79 - 1.66) | 0.70<br>(0.40 - 1.23) | 1.81<br>(0.94 - 3.47) |
| Education: US: bachelor or higher /<br>DE: Abitur or technical diploma = 1 | 0.99<br>(0.68 - 1.44) | 1.26<br>(0.86 - 1.85) | 0.78<br>(0.41 - 1.45) | 0.65<br>(0.32 - 1.29) |
| Employment: employed (any type) = 1 | 0.91<br>(0.63 - 1.32) | 0.76<br>(0.48 - 1.20) | 0.89<br>(0.49 - 1.61) | 0.94<br>(0.46 - 1.95) |
| Been vaccinated = 1 | 1.28<br>(0.88 - 1.86) | 0.85<br>(0.57 - 1.26) | 1.32<br>(0.73 - 2.38) | 1.49<br>(0.76 - 2.95) |
| SD5 | 1.00<br>(0.99 - 1.00) | 1.00<br>(0.99 - 1.01) | 1.00<br>(0.99 - 1.01) | 1.02**<br>(1.00 - 1.03) |
| Life affected by COVID-19 | 0.97<br>(0.88 - 1.06) | 1.02<br>(0.93 - 1.13) | 1.03<br>(0.89 - 1.21) | 1.06<br>(0.90 - 1.24) |
| Constant | 0.27***<br>(0.16 - 0.44) | 0.19***<br>(0.10 - 0.36) | 0.07***<br>(0.03 - 0.15) | 0.02***<br>(0.01 - 0.06) |
| Observations | 1,000 | 1,003 | 1,000 | 1,003 |

Robust CI (eform) in parentheses; \*\*\* $p < 0.01$ , \*\* $p < 0.05$ . Age equal to or greater than 70 = 1, otherwise 0; education level in the United States (US) as “bachelor degree or higher” = 1 and in as Germany (DE), “abitur / technical diploma” = 1, otherwise 0; employment status as “employed (any type)” = 1, otherwise 0.

Figure 2 illustrates the extent to which respondents were willing to wait three more months for their own vaccine so that people in countries with certain characteristics could receive it earlier (7-point scale, 1 = *Strongly disagree* to 7 = *Strongly agree*). This question was particularly critical at the time when many respondents were still waiting to be vaccinated themselves during the survey in June 2021. In both the United States and Germany the highest level of agreement was with waiting so that people in countries with higher number of COVID-19 deaths followed by those with fewer ICU beds could be vaccinated earlier.

**Figure 2.**
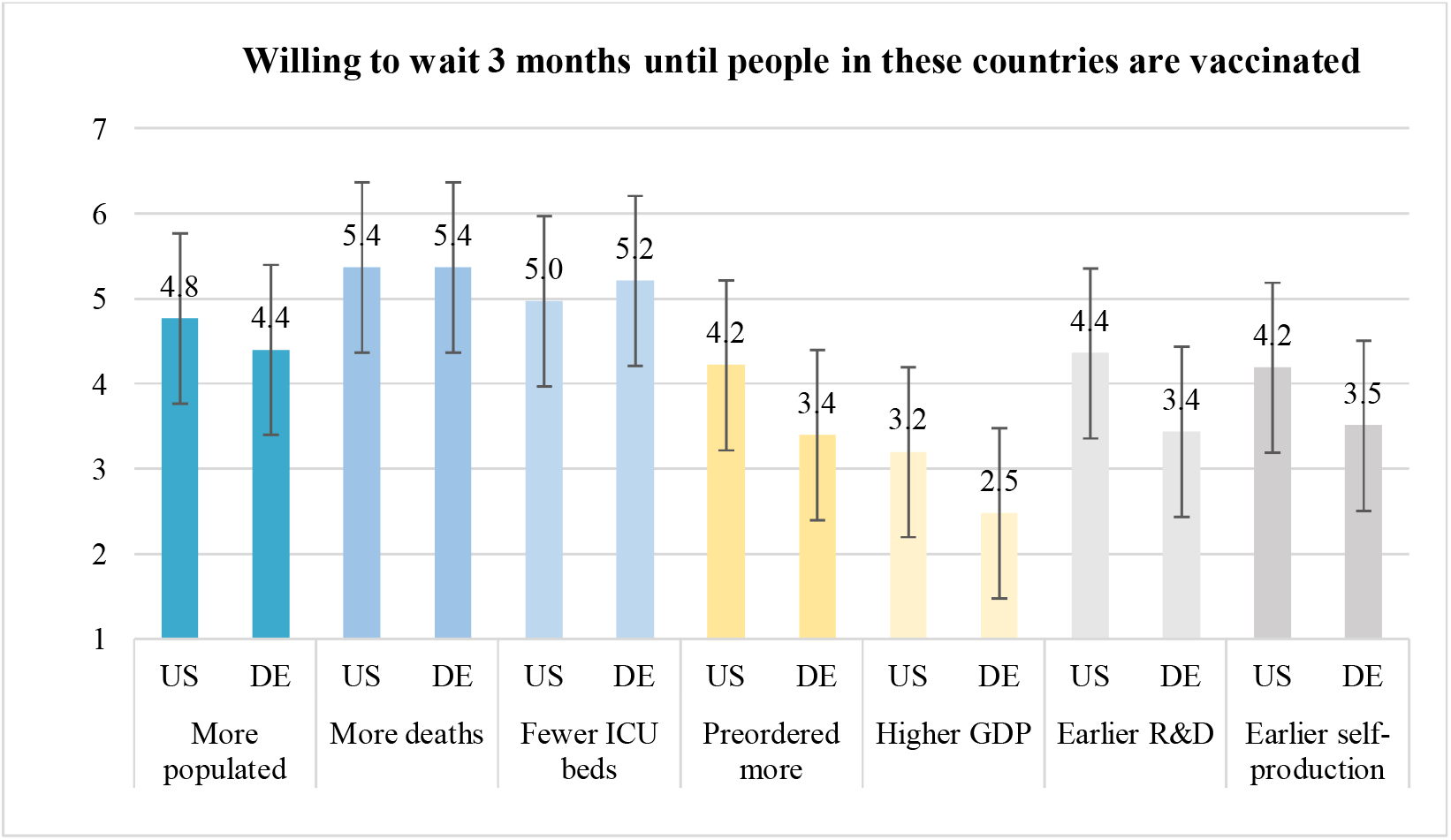
Mean level of willingness to wait on a 7-point scale (1 = *Strongly disagree* – 7 = *Strongly agree*).

## Discussion

Our study measured public attitude towards fair global distribution of COVID-19 vaccines. Using an analytical hierarchy process, we asked respondents to weigh various principles that could guide the distribution of COVID-19 vaccines across the world. We found that the general public in both, the United States and Germany, gave the highest weighting to principles that correspond with utilitarian values followed by egalitarian values. Despite both countries being pioneers in research and development of the vaccines and also having own production capacities, the public in neither of these countries gave a high weighting to principles that correspond with merit based considerations. As it stands, however, access to vaccines is ruled by free market principles – which in this study was given the lowest weighting by the general public. In a discrete choice experiment we asked respondents to divide scarce vaccination resources between a high- and a low-income country. When we changed the scenario such that a vulnerable family member or the respondents themselves were affected by this decision, only a relatively small share of respondents reduced the amount that was given to the other country by enough to ensure sufficient supply for their vulnerable family member or themselves. The decision to reduce the amount given to the low-income country was not correlated with the respondent’s gender, education level, employment status, vaccination status or negative effects of COVID-19 on their life. And there was only a weak correlation with old age. Based on the Marlowe-Crowne scale we did not find that any of the results were driven by social desirability bias. However, our study has the limitation of reporting a cross-sectional observation of a dynamic and rapidly changing topic.

The presented results connect strongly to current discussions initiated – among others – by multilateral organizations like the IMF, the World Bank and the WHO. As for example presented in the World Economic Outlook, global economic recovery hinges very strongly on vaccine distribution issues; the current distribution imbalances are found to trigger severe economic imbalances in the future (20). From a public health angle, the WHO is raising attention towards growing disparities as HICs proceed towards third booster vaccine shots, whereas many LMICs are still struggling to access and distribute first and second COVID-19 vaccine doses ((21), p. 63). If we think of humanity as one population, we are steering towards a situation where a large share of the population is vaccinated and a large share is not. This is a critical phase of the pandemic because it provides ideal conditions for the virus SARS-Cov-2 to escape the vaccine and to emerge stronger than before through mutation and virus variants (22). Therefore, the pandemic will not be over before it is over everywhere on the globe. Public perceptions regarding distribution of COVID-19 vaccine doses that we report in this study are very much in line with this view of humanity as one population – which is both, an expression of fairness and arguably the best way out of the pandemic. Unfortunately, the actions of HICs are not matching this expressed opinion of the general public. We are now in a situation where there are more vaccine doses available than people willing to be vaccinated in HICs – and the opposite in LMICs (23); which is both, short-sighted and against the preferences of the general public. Aligning public action in HICs with these public perceptions might go a long way in overcoming the pandemic and valuing the view of the people at the same time.

## Data Availability

The data will be available after the successful dissemination of the study results.

## Acknowledgments

Not applicable.

## Ethics committee approval

This study received ethical approval from the ethics committee at the University of Göttingen, Germany.

## Abbreviations

AHP: Analytical hierarchy process
COVAX: the COVID-19 Vaccine Global Access program
GDP: Gross domestic product
HICs: High-income countries
ICU: Intensive care unit
LMICs: Low- and middle-income countries
(OR): Odds ratio
R&D: Research and development
SD: Social desirability.

## References

1. Buheji M, da Costa Cunha K, Beka G, Mavrić B, Leandro do Carmo de Souza Y, Da Souza Costa Silva S et al. The Extent of COVID-19 Pandemic Socio-Economic Impact on Global Poverty. A Global Integrative Multidisciplinary Review. economics 2020; 10(4):213–24.

2. Cohen J. Countries now scrambling for COVID-19 vaccines may soon have surpluses to donate. Science 2021.

3. Herzog LM, Norheim OF, Emanuel EJ, McCoy MS. Covax must go beyond proportional allocation of covid vaccines to ensure fair and equitable access. The BMJ 2021; 372.

4. Emanuel EJ, Persad G, Kern A, Buchanan A, Fabre C, Halliday D et al. An ethical framework for global vaccine allocation. Science 2020; 369(6509):1309–12.

5. Our World in Data. Coronavirus (COVID-19) Vaccinations; 2021. Available from: URL: https://ourworldindata.org/covid-vaccinations. Last accessed 03/11/2021.

6. International Monetary Fund. IMF-WHO COVID-19 Vaccine Supply Tracker [cited 10/30/2021]. Available from: URL: https://www.imf.org/en/Topics/imf-and-covid19/IMF-WHO-COVID-19-Vaccine-Supply-Tracker. Last accessed 03/11/2021.

7. Fontanet A, Autran B, Lina B, Kieny MP, Karim SS, Sridhar D. SARS-CoV-2 variants and ending the COVID-19 pandemic. The Lancet 2021; 397(10278):952–4.

8. Wouters OJ, Shadlen KC, Salcher-Konrad M, Pollard AJ, Larson HJ, Teerawattananon Y et al. Challenges in ensuring global access to COVID-19 vaccines: production, affordability, allocation, and deployment. The Lancet 2021; 397(10278):1023–34.

9. Herlitz A, Lederman Z, Miller J, Fleurbaey M, Venkatapuram S, Atuire C et al. Just allocation of COVID-19 vaccines. BMJ Glob Health 2021; 6(2):e004812.

10. Usher AD. A beautiful idea: how COVAX has fallen short. The Lancet 2021; 397(10292):2322–5.

11. Conybeare JAC. Efficiency, entitlements and deservingness: Perspectives on international distributive justice. Review of International Political Economy 2007; 14(3):389–411.

12. Stilz A. Is The Free Market Fairã Critical Review 2014; 26(3-4):423–38.

13. Guidry JPD, Perrin PB, Laestadius LI, Vraga EK, Miller CA, Fuemmeler BF et al. U.S. public support for COVID-19 vaccine donation to low- and middle-income countries during the COVID-19 pandemic. Vaccine 2021; 39(17):2452–7.

14. Clarke PM, Roope LSJ, Loewen PJ, Bonnefon J-F, Melegaro A, Friedman J et al. Public opinion on global rollout of COVID-19 vaccines. Nat Med 2021; 27(6):935–6.

15. Our World in Data. Daily new confirmed COVID-19 deaths per million people; 2021 [cited 2021 Oct 5]. Available from: URL: https://ourworldindata.org/explorers/coronavirus-data-explorer?zoomToSelection=true&facet=none&pickerSort=desc&pickerMetric=location&Interval=7-day+rolling+average&Relative+to+Population=true&Align+outbreaks=false&country=USA~DEU~OWID_WRL&Metric=Confirmed+deaths. Last accessed 03/11/2021.

16. Huang K, Greene JD, Bazerman M. Veil-of-ignorance reasoning favors the greater good. Proc Natl Acad Sci USA 2019; 116(48):23989–95.

17. Huang K, Bernhard R, Barak-Corren N, Bazerman M, Greene JD. Veil-of-Ignorance Reasoning Mitigates Self-Serving Bias in Resource Allocation During the COVID-19 Crisis. Center for Open Science; 2020.

18. Hays RD, Hayashi T, Stewart AL. A five-item measure of socially desirable response set. Educational and Psychological Measurement 1989; 49(3):629–36.

19. Steptoe A, Deaton A, Stone AA. Subjective wellbeing, health, and ageing. The Lancet 2015; 385(9968):640–8.

20. Gita Gopinath. IMF World Economic Outlook July 2021: Vaccine differences drive economic differences. Available from: URL: https://blogs.imf.org/2021/07/27/drawing-further-apart-widening-gaps-in-the-global-recovery/. Last accessed 03/11/2021.

21. The Independent Panel on Pandemic Preparedness and Response. COVID-19: COVID-19: Make it the Last Pandemic. GENEVA; 2021. Available from: URL: https://theindependentpanel.org/wp-content/uploads/2021/05/COVID-19-Make-it-the-Last-Pandemic_final.pdf. Last accessed 03/11/2021.

22. Mascola JR, Graham BS, Fauci AS. SARS-CoV-2 viral variants - tackling a moving target. JAMA 2021; 325(13):1261–2.

23. Solís Arce JS, Warren SS, Meriggi NF, Scacco A, McMurry N, Voors M et al. COVID-19 vaccine acceptance and hesitancy in low- and middle-income countries. Nat Med 2021; 27:1385–94.

