## supplementary material for "Public opinion on global distribution of COVID-19 vaccines: evidence from two nationally representative surveys in Germany and the United States"

**Appendix A: Supplementary material for the manuscript “Public opinion on global distribution of COVID-19 vaccines: evidence from two nationally representative surveys in Germany and the United States”**

**Section I: Further results**

Table A1 presents the respondents demographics in the United States (US) and Germany (DE).

| **Table A1.** Demographics of survey participants (weighted). | | | | | | | | | |
| --- | --- | --- | --- | --- | --- | --- | --- | --- | --- |
|  | US (*N* = 1000) | | | | | DE (*N* = 1003) | | | |
|  | Mean | Sd. | Min | Max | Mean | | Sd. | Min | Max |
| Age | 47·4 | 17·8 | 18 | 90 | 49·8 | | 17·2 | 18 | 91 |
| Female (%) | 51·3 |  |  |  | 51·4 | |  |  |  |
| Been vaccinated (%) | 64·8 |  |  |  | 60·9 | |  |  |  |
| Education: US / DE (%) |  |  |  |  |  | |  |  |  |
| Less than 9^th^ grade / Up to 8 classes | 1·1 |  |  |  | 3·8 | |  |  |  |
| 12^th^ grade; no diploma /9^th^ grade; secondary | 6·2 |  |  |  | 16·6 | |  |  |  |
| High school / 10^th^ grade; middle school | 57·9 |  |  |  | 48·4 | |  |  |  |
| Bachelor's degree; higher / Abitur; technical diploma | 30·6 |  |  |  | 30·6 | |  |  |  |
| No degree | 4·1 |  |  |  | 0·7 | |  |  |  |
| Employment (%) |  |  |  |  |  | |  |  |  |
| Full-time | 33·1 |  |  |  | 39·7 | |  |  |  |
| Self-employed full-time | 4·3 |  |  |  | 4·3 | |  |  |  |
| Part-time | 10·4 |  |  |  | 7·8 | |  |  |  |
| Part-time (want full-time) | 2·8 |  |  |  | 1·3 | |  |  |  |
| Mini job (up to 450 euros) | *NA* |  |  |  | 4 | |  |  |  |
| Seeking employment | 13·7 |  |  |  | 3·5 | |  |  |  |
| Without employment | 35·7 |  |  |  | 39·4 | |  |  |  |

**Section II: Survey instrument (English version)**

**Global Distribution of COVID 19 Vaccines**

This is a survey aiming to study the public opinion on the distribution of COVID-19 vaccination across the world. The survey has 14 questions and it should take around 10 to 15 minutes to complete. Your response will be anonymized; the data will remain protected and treated with utmost confidentiality. This study is for academic purposes and is conducted by a team of researchers at the Georg-August-University of Göttingen in Germany. Your help will be highly appreciated. Would you be willing to participate?

**Consent**

Could you please confirm that you are an adult (aged 18 or older) and are willing to participate in this survey?

- Yes
- No *(end the questionnaire).*

### A. General Information

| 1. Could you please state your **year of birth**? | \|___\| \|___\| \|___\| \|___\| |
| --- | --- |
| 2. How would you describe your **gender**: | - Male (including transgender men) - Female (including transgender women) - Prefer to self-describe as (non-binary, gender-fluid, gender, please specify) (free-text) _____________ |
| 3. Please provide us with the first three digits of your **postcode** | \|___\| \|___\| \|___\| |
| 4. What is the state of your employment: | - Employed full time for an employer (work for at least 30 hours per week) - Employed full time for self (work for at least 30 hours per week) - Employed part time (by self or other) and do not want to work full time (work less than 30 hours per week) - Employed part time, want to work full time - Unemployed - Out of the workforce |

### B. Opinion I

The following questions will ask you about your opinion on how COVID-19 vaccines should be distributed across the world. Therefore, we discuss countries and not individual persons. Please note that there are no "right" or "wrong" answer to these questions.

| 5. Think about the distribution of COVID-19 vaccines across the world and possible **principles to allocate vaccine shipments**. Please compare two principles each and rate the importance within these pairs.  *(Show only one comparison at a time, random order, implement “slider” online)* | | | | | | | | | | | | | | | | | | |
| --- | --- | --- | --- | --- | --- | --- | --- | --- | --- | --- | --- | --- | --- | --- | --- | --- | --- | --- |
|  | 9 | 8 | 7 | 6 | 5 | 4 | 3 | 2 | 1 | 2 | 3 | 4 | 5 | 6 | 7 | 8 | 9 |  |
| **A** | Extremely Important |  |  |  |  |  |  |  | Equally Important |  |  |  |  |  |  |  | Extremely Important | **B** |
| Equal Access for All ^a^ |  |  |  |  |  |  |  |  |  |  |  |  |  |  |  |  |  | Medical Urgency ^b^ |
| Equal Access for All |  |  |  |  |  |  |  |  |  |  |  |  |  |  |  |  |  | Free Market Rules Apply ^c^ |
| Equal Access for All |  |  |  |  |  |  |  |  |  |  |  |  |  |  |  |  |  | Production Contribution ^d^ |
| Medical Urgency |  |  |  |  |  |  |  |  |  |  |  |  |  |  |  |  |  | Free Market Rules Apply |
| Medical Urgency |  |  |  |  |  |  |  |  |  |  |  |  |  |  |  |  |  | Production Contribution |
| Free Market Rules Apply |  |  |  |  |  |  |  |  |  |  |  |  |  |  |  |  |  | Production Contribution |

**Hints (Mouse-over Info):*

*^a^ Equal Access for All: is based on the principle of “one person one vote” and everyone should be treated equally regardless of their individual conditions.*

*^b^ Medical Urgency: is based on the principle that distribution should be according to benefits and needs in a medical sense given health systems and personal medical requirements.*

*^c^ Free Market Rules Apply: is based on the principle that the market is a fair and free playfield and seller’s and customer’s rights are protected as long as both sides are happy with their exchange.*

*^d^ Production Contribution: is based on merits rewarding those who have invested in the research and development of the vaccines and/or those who have the capacity to produce vaccines in their own country.*

| 6. Suppose that there are **three vaccine production and distribution steps** upcoming:  -- **Today**, 100 million vaccine doses are available for the two countries described below.  -- In about **six months**, there will be an additional 100 million doses ready for distribution.  -- In about **12 months**, there will be enough doses for vaccinating all people.  Assume that one dose is sufficient to vaccinate one person. **Please** decide about the vaccine distribution in the first two steps between the two countries: *(the sum of each row should add up to 100 Mil.)* | |
| --- | --- |
| **Country A** | **Country B** |
| - *100 million inhabitants* - *200 COVID-19 deaths per day* - *50 intensive care hospital beds (per 100,000)* - *1,000 million ordered vaccine doses* - *75,000 US-$ average yearly income* - *Completed vaccine research investment* - *Vaccine production capacity* | - *300 million inhabitants* - *3,000 COVID-19 deaths per day* - *4 intensive care hospital beds (per 100,000)* - *100 million ordered vaccine doses* - *7,000 US-$ average yearly income* - *No previous vaccine investment* - *No own vaccine production capacity* |
| \|_______\| Mil. doses | \|_______\| Mil. doses |
| 7a. Would you like to explain why you made these decisions and what were your line of thoughts:  ____________________________________________________________________________  ____________________________________________________________________________ | |

| 7. Regarding COVID-19 vaccination, you are now thinking of a **close family member of old age** with a higher risk due to COVID-19 living with you in **Country A**. This person you care a lot about received the information to be being the X millionth person in line for vaccination [X= Q6 Entry Country A + 10 Million].  Again, there are **three vaccine production and distribution steps** upcoming for shipments into the two countries below:  **[1]** **Today**, 100 million vaccine doses are available for the two countries described below.  **[2]** In about **six months**, there will be an additional 100 million doses ready for distribution.  **[3]** In about **12 months**, there will be enough doses for vaccinating all people.  Again, assume that one dose is sufficient to vaccinate one person. Please decide about the vaccine distribution in the first two steps between the two countries:  *(the sum of each row should add up to 100 Mil.)* | |
| --- | --- |
| **Country A** | **Country B** |
| - *100 million inhabitants* - *200 COVID-19 deaths per day* - *50 intensive care hospital beds (per 100,000)* - *1,000 million ordered vaccine doses* - *75,000 US-$ average yearly income* - *Completed vaccine research investment* - *Vaccine production capacity* | - *300 million inhabitants* - *3,000 COVID-19 deaths per day* - *4 intensive care hospital beds (per 100,000)* - *100 million ordered vaccine doses* - *7,000 US-$ average yearly income* - *No previous vaccine investment* - *No own vaccine production capacity* |
| \|_______\| Mil. doses | \|_______\| Mil. doses |
| 8a. Would you like to explain why you made these decisions and what were your line of thoughts:  ___________________________________________________________________________  ___________________________________________________________________________ | |

| 8. **You** have received the information to be the X millionth person in line in **Country A** [X= Q6 Entry Country A + 30 Million].  Again, there are **three vaccine production and distribution steps** upcoming for shipments into the two countries below:  **[1]** **Today**, 100 million vaccine doses are available for the two countries described below.  **[2]** In about **six months**, there will be an additional 100 million doses ready for distribution.  **[3]** In about **12 months**, there will be enough doses for vaccinating all people.  Again, assume that one dose is sufficient to vaccinate one person. Please decide about the vaccine distribution in the first two steps between the two countries:  *(the sum of each row should add up to 100 Mil.)* | |
| --- | --- |
| **Country A** | **Country B** |
| - *100 million inhabitants* - *200 COVID-19 deaths per day* - *50 intensive care hospital beds (per 100,000)* - *1,000 million ordered vaccine doses* - *75,000 US-$ average yearly income* - *Completed vaccine research investment* - *Vaccine production capacity* | - *300 million inhabitants* - *3,000 COVID-19 deaths per day* - *4 intensive care hospital beds (per 100,000)* - *100 million ordered vaccine doses* - *7,000 US-$ average yearly income* - *No previous vaccine investment* - *No own vaccine production capacity* |
| \|_______\| Mil. doses | \|_______\| Mil. doses |
| 9a. Would you like to explain why you made these decisions and what were your line of thoughts:  ___________________________________________________________________________  ___________________________________________________________________________ | |

| 9. To what extent do you agree or disagree with the following statements? | | | | | | | |
| --- | --- | --- | --- | --- | --- | --- | --- |
| *(random order)* | *Strongly disagree* | *Disagree* | *Somewhat disagree* | *Neither agree nor disagree* | *Somewhat agree* | *Agree* | *Strongly Agree* |
| *Example* |  |  |  |  | **X** |  |  |
| **I would be willing to wait three months longer for my own vaccination …** | | | | | | | |
| … in order to vaccinate people in more populated countries earlier. |  |  |  |  |  |  |  |
| … in order to vaccinate people in countries with higher number of COVID-19 deaths earlier. |  |  |  |  |  |  |  |
| … in order to vaccinate people in countries with a lower number of intensive care hospital beds (per 100,000 inhabitants) earlier. |  |  |  |  |  |  |  |
| … in order to vaccinate people in countries with a higher number of vaccine pre-orders. |  |  |  |  |  |  |  |
| … in order to vaccinate people in countries with a higher annual income per head earlier. |  |  |  |  |  |  |  |
| … in order to vaccinate people in countries who have invested in research and development of the vaccines earlier. |  |  |  |  |  |  |  |
| … in order to vaccinate people in countries who are able to produce vaccines in their country earlier. |  |  |  |  |  |  |  |

| 10. To what extent do you agree or disagree with the following statements? | | | | | | |
| --- | --- | --- | --- | --- | --- | --- |
| *(random order)* | *Definitely false* | *Mostly false* | | *Don't know* | *Mostly true* | *Definitely true* |
| *Example* |  |  |  |  | **X** |  |
| I am always courteous even to people who are disagreeable. |  |  | |  |  |  |
| There have been occasions when I took advantage of someone. |  |  | |  |  |  |
| I sometimes try to get even rather than forgive and forget. |  |  | |  |  |  |
| I sometimes feel resentful when I don’t get my way. |  |  | |  |  |  |
| No matter who I’m talking to, I’m always a good listener. |  |  | |  |  |  |

### C. Personal Information

| 11. To what extent do you agree or disagree with the following statement? | | | | | | | |
| --- | --- | --- | --- | --- | --- | --- | --- |
|  | *Strongly disagree* | *Disagree* | *Somewhat disagree* | *Neither agree nor disagree* | *Somewhat agree* | *Agree* | *Strongly Agree* |
| *Example* |  |  |  |  | **X** |  |  |
| The COVID-19 pandemic has had a strong negative impact on my (or my loved-ones’) life (s). |  |  |  |  |  |  |  |

| 12. Have you already been vaccinated against COVID-19 or do you have an appointment to be vaccinated? | - Yes - No |
| --- | --- |

### D. Opinion II

| 13. Think about the distribution of COVID-19 vaccines across the world to **specific countries** and possible **criteria to allocate vaccine shipments**. Please **compare two criteria** each and rate the importance within these pairs. *(Show only one comparison at a time, random order, implement “slider” online)* | | | | | | | | | | | | | | | | | | |
| --- | --- | --- | --- | --- | --- | --- | --- | --- | --- | --- | --- | --- | --- | --- | --- | --- | --- | --- |
|  | 9 | 8 | 7 | 6 | 5 | 4 | 3 | 2 | 1 | 2 | 3 | 4 | 5 | 6 | 7 | 8 | 9 |  |
| **A** | Extremely Important |  |  |  |  |  |  |  | Equally Important |  |  |  |  |  |  |  | Extremely Important | **B** |
| Number of inhabitants |  |  |  |  |  |  |  |  |  |  |  |  |  |  |  |  |  | Number of daily COVID-19 deaths |
| Number of inhabitants |  |  |  |  |  |  |  |  |  |  |  |  |  |  |  |  |  | Number of intensive care hospital beds (per 100.000) |
| Number of inhabitants |  |  |  |  |  |  |  |  |  |  |  |  |  |  |  |  |  | Number of vaccines pre-ordered |
| Number of inhabitants |  |  |  |  |  |  |  |  |  |  |  |  |  |  |  |  |  | Annual income per head (GDP) |
| Number of inhabitants |  |  |  |  |  |  |  |  |  |  |  |  |  |  |  |  |  | Invested in vaccine research and development |
| Number of inhabitants |  |  |  |  |  |  |  |  |  |  |  |  |  |  |  |  |  | Vaccine production capacity |
| Number of daily COVID-19 deaths |  |  |  |  |  |  |  |  |  |  |  |  |  |  |  |  |  | Number of intensive care hospital beds (per 100.000) |
| Number of daily COVID-19 deaths |  |  |  |  |  |  |  |  |  |  |  |  |  |  |  |  |  | Number of vaccines pre-ordered |
| Number of daily COVID-19 deaths |  |  |  |  |  |  |  |  |  |  |  |  |  |  |  |  |  | Annual income per head (GDP) |
| Number of daily COVID-19 deaths |  |  |  |  |  |  |  |  |  |  |  |  |  |  |  |  |  | Invested in vaccine research and development |
| Number of daily COVID-19 deaths |  |  |  |  |  |  |  |  |  |  |  |  |  |  |  |  |  | Vaccine production capacity |
| Number of intensive care hospital beds (per 100.000) |  |  |  |  |  |  |  |  |  |  |  |  |  |  |  |  |  | Number of vaccines pre-ordered |
| Number of intensive care hospital beds (per 100.000) |  |  |  |  |  |  |  |  |  |  |  |  |  |  |  |  |  | Annual income per head (GDP) |
| Number of intensive care hospital beds (per 100.000) |  |  |  |  |  |  |  |  |  |  |  |  |  |  |  |  |  | Invested in vaccine research and development |
| Number of intensive care hospital beds (per 100.000) |  |  |  |  |  |  |  |  |  |  |  |  |  |  |  |  |  | Vaccine production capacity |
| Number of vaccines pre-ordered |  |  |  |  |  |  |  |  |  |  |  |  |  |  |  |  |  | Annual income per head (GDP) |
| Number of vaccines pre-ordered |  |  |  |  |  |  |  |  |  |  |  |  |  |  |  |  |  | Invested in vaccine research and development |
| Number of vaccines pre-ordered |  |  |  |  |  |  |  |  |  |  |  |  |  |  |  |  |  | Vaccine production capacity |
| Annual income per head (GDP) |  |  |  |  |  |  |  |  |  |  |  |  |  |  |  |  |  | Invested in vaccine research and development |
| Annual income per head (GDP) |  |  |  |  |  |  |  |  |  |  |  |  |  |  |  |  |  | Vaccine production capacity |
| Invested in vaccine research and development |  |  |  |  |  |  |  |  |  |  |  |  |  |  |  |  |  | Vaccine production capacity |

Option: Would you like to leave any further comments?

*Closing statement: We have reached to the end of the survey; thank you so much for your participation. If you have any questions regarding this survey please contact* *.*
